# Rapid Detection of SARS-CoV-2 Using Reverse transcription RT-LAMP method

**DOI:** 10.1101/2020.03.02.20030130

**Authors:** Weihua Yang, Xiaofei Dang, Qingxi Wang, Mingjie Xu, Qianqian Zhao, Yunying Zhou, Huailong Zhao, Li Wang, Yihui Xu, Jun Wang, Shuyi Han, Min Wang, Fenyan Pei, Yunshan Wan

## Abstract

Corona Virus Disease 2019 (COVID-19) is a recently emerged life-threatening disease caused by SARS-CoV-2. Real-time fluorescent PCR (RT-PCR) is the clinical standard for SARS-CoV-2 nucleic acid detection. To detect SARS-CoV-2 early and control the disease spreading on time, a faster and more convenient method for SARS-CoV-2 nucleic acid detecting, RT-LAMP method (reverse transcription loop-mediated isothermal amplification) was developed. RNA reverse transcription and nucleic acid amplification were performed in one step at 63 °C isothermal conditions, and the results can be obtained within 30 minutes. ORF1ab gene, E gene and N gene were detected at the same time. ORF1ab gene was very specific and N gene was very sensitivity, so they can guarantee both sensitivity and specificity for SARS-CoV-2. The sensitivity of RT-LAMP assay is similar to RT-PCR, and specificity was 99% as detecting 208 clinical specimens. The RT-LAMP assay reported here has the advantages of rapid amplification, simple operation, and easy detection, which is useful for the rapid and reliable clinical diagnosis of SARS-CoV-2.

## Introduction

Corona Virus Disease 2019 (COVID-19) is a recently emerged life-threatening viral pneumonia disease caused by SARS-CoV-2 (named 2019-nCoV before February 11, 2020)(1). COVID-19 is spreading worldwide, and as of February 29, 2020, approximately 80,000 confirmed cases had been identified in the world. SARS-CoV-2 belongs to the beta coronavirus genus and its genome is single-stranded, non-segmented positive-sense RNA, which is 29,881 nucleotides in length(2). It has 84% nucleotide homology with bat SARS-like coronavirus, 78% homology with human SARS virus, and approximately 50% homology with MERS virus(3, 4). So far, the full-length genomic sequences between virus samples are almost identical(3), suggesting that no significant mutations have occurred in the virus(5).

There is no suitable antiviral drug or effective vaccine for the SARS-CoV-2 virus currently. There may be cross-reactions in serum antibody test (6). Therefore, reliable and rapid laboratory diagnosis of SARS-CoV-2 infection is critical to managing patient care and preventing hospital transmission. For clinical diagnosis of COVID-19, nucleic acid detection or sequencing is currently used in conjunction with pulmonary CT(7). Real-time reverse-transcription fluorescent PCR (RT-PCR) is the clinical standard for SARS-CoV-2 nucleic acid detection(8). Now, in order to detect SARS-CoV-2 early and control the disease spreading on time, we developed a faster and more convenient method for detecting SARS-CoV-2 nucleic acid, named RT-LAMP (reverse transcription loop-mediated isothermal amplification). ORF1ab gene, E gene and N gene were selected and primers for them were designed. We have used the RT-LAMP method to detect pathogenic microorganisms and achieved good results(9, 10). In 2003, RT-LAMP had been reported as a quick and easy diagnostic method for SARS coronavirus(11, 12).

LAMP is a kind of isothermal amplification method that can amplify nucleic acid with high specificity, sensitivity, and rapidity at 60-65 °C and does not require special instruments such as a thermal cycler. The amplification efficiency of the LAMP method is extremely high, the reaction can be completed within one hour. This method relies on self-recycling strand displacement DNA synthesis by Bst DNA polymerase, and previous studies have described detailed amplification mechanisms(13). This reaction relies on the recognition of DNA targets by six independent sequences, making this assay highly specific. When the template is RNA, the amplification reaction can be occurred in one step by adding reverse transcriptase, and called RT-LAMP.

Magnesium pyrophosphate is produced as a by-product during DNA amplification. Because the amplification efficiency of LAMP is higher than that of RT-PCR, a large amount of magnesium pyrophosphate causes the reaction solution to become cloudy. Besides, the added dye calcein can change the mixed solution from orange to green, so the results can not only be observed by a turbidimeter, but also by the naked eye. Because RT-LAMP is convenient and fast, it shows significant advantages in SARS-CoV-2 detection in clinic(14). Sensitivity, specificity and application potential of RT-LAMP in the clinical diagnosis of SARS-CoV-2 were reported and discussed here.

## Materials and methods

### Clinical samples

Our hospital is one of the six hospitals for SARS-CoV-2 nucleic acid testing approved by our city. We have five positive specimens and more than 4,000 negative specimens. Positive samples from our hospital were used for the method developing. Specimens from our hospital, Infectious disease hospital of Jinan and Jinan Center for Disease Control and Prevention (Jinan CDC) were used in the clinical verification stage. From January 30 to February 27, during the SARS-CoV-2 epidemic, a total of 208 RNA from throat swab specimens were collected and 17 were SARS-CoV-2 positive.

### RNA extraction

RNA was extracted using Liferiver (Shanghai Liferiver Biotechnology Co., Ltd.) nucleic acid extraction kit by Nucleic acid extraction instrumentin a tertiary protection laboratory.

### Designing of primers

In order to identify SARS-CoV-2, the species-specific ORF1ab gene, E gene and N gene were selected as the target gene (GenBank accession No.MN988669.1) after comparing the gene sequences of CoVs from human and bat origins. Oligonucleotide primers for LAMP were designed using Primer Explorer V5 software (http://primerexplorer.jp/e/), and synthesized commercially by invitrogen biotech (Invitrogen, Carlsbad, CA, USA). All primers were compared with other coronaviruses and published SARS-CoV-2 sequences by genebank using BLAST alignment tools. Results showed the sequences of the selected primers were specific. Ten sets primers for ORF1ab gene, 12 sets primers for E gene and 13 sets primers for N were designed and synthesized. Each set composed by four primers (F3, forward outer primer; B3, backward outer primer; FIP, forward inner primer; BIP, backward inner primer) targeting six distinct regions (Tsugunori, 2000). After the suitable primer sets were screened out, loop primers (LF/LP) of them were designed to accelerate the amplification.

### Primer Screening

Primer screening is divided into three steps. The first step is to screen primers that can react. The primers were screened using two positive samples, and the primers that could react within 60 minutes were selected. The second step is to screen primers without false positives reaction. Ten negative samples were used for the reaction. The primers without false positive reactions were selected and others were discarded. The third step is to screen the loop primers. Loop primers of the selected primers were designed. Then the loop primers which can react within 30 minutes were selected.

### RT-LAMP Reaction

RT-LAMP was carried out using a Loopamp reverse transcription nucleic acid amplification kit (Eiken China Co., Ltd., Shanghai, China) according to the manufacturers’ instructions. Brifely, a 25µL total reaction mixture containing 12.5 µL of 2×Reaction Mix [Tris-HCl 40 mM (pH 8.8), KCl 20 mM, MgSO_4_ 16 mM, (HN4)_2_SO_4_ 20 mM, Tween20 0.2%, betain 1.6 M, dNTPs 2.8 mM*4], 1.0 µL Enzyme solution (8 U Bst DNA polymerase, 200 U AMV Reverse transcriptase), 1.0 µL Fluorescent Detection Reagent (Eiken),1.0 µL each of the outer primers (5 pmol), 1.0 µL each of the inner primers (40 pmol), 1.0 µL loop primers (20 pmol), 2.0 µL of target RNA template and 2.5/3.5µL DEPC treated water were added together to the reaction tube. Negative and positive controls were established according to the kit instructions. And then the reaction tubes were incubated at 63 °C for 60 minutes in a Loopamp real-time turbidimeter (LA-320 C;Eiken Chemical Co., Ltd. Tokyo, Japan) to screen out the optimal primer sets.

### RT-PCR Reaction

All the samples we used had been tested by a RT-PCR kit (Jiangsu Bioperfectus Technologies Co.,Ltd.) and confirmed by clinical symptoms and epidemiology of patients. The kit had been registered and it detects ORF1ab gene, E gene and N gene too. RT-PCR was carried out according to the manufacturers’ instruction. Briefly, a 25 µL total reaction mixture containing 19µL detection mixture, 1µL RT-PCR enzyme and 5 µL target RNA template were mixed in a reaction tube. Then RT-PCR was performed (45 °C for 10 min, 95 °C for 3 min, 95 °C for 15 sec,58 °C for 30 sec, 45 cycles) in ABI 7500 Real-time PCR system (Applied Biosystems, USA).

### Interpretation of RT-LAMP results

The real-time amplification by RT-LAMP assay was monitored through spectrophotometric analysis by recording the optical density at 400 nm every 6 s with the help of the Loopamp real-time turbidimeter. Besides,the LAMP products can be detected based on color change. The color of the reaction mixture turned green in the presence of LAMP amplification, while the color remained orange in negative amplification tubes. RT-LAMP products were 2-fold serial diluted, and then observed electrophoresed. Because the amount of amplification products is very large, aerosols will be formed when open the reaction tube cover and will pollute the environment. It is not recommended to open the cover and observe results by electrophoresis. Electrophoresis was not used as a way to judge results no matter when, and here, we just for illustration.

### Determination of reaction time

As reaction time increases, false positives will appear. In order to determine the timing for judging results, five positive samples and ten negative samples were tested, and the performance of three genes at different time points was compared. When five samples were positive for all three genes, and false positives did not appear, was the time to judge the results.

### Sensitivity of RT-LAMP assays

To determine the sensitivity of the RT-LAMP method using the screened primers to identify ORF1ab gene, E gene and N gene of SARS-CoV-2, RNA No. 101 was serially 2-fold diluted, ranging from 10,20,40,80 to 160 folds, RT-LAMP and RT-PCR were performed at the same time using these diluted RNA. The minimum concentration of their positive reaction was observed and recorded. The sensitivity of RT-LAMP was determined by comparison with PCR. The detection limit of the PCR kit used is 1000 copies/ml.

### Specificity of RT-LAMP identify

To validate the specificity of the RT-LAMP assays, RNA of 10 Influenza A, Influenza B, Respiratory Syncytial Virus positive samples and 20 SARS-CoV-2 negative samples was amplified by RT-LAMP as mentioned above. The RT-PCR method was used for comparison. The RT-LAMP products were assayed by 3% agarose gel electrophoresis and visualized under UVI gel-image analysis system (UVI, UK). DNA fragments were recovered by gel cutting, and the specificity of the fragments was verified by sequencing.

### Interference test

During the outbreak of SARS-CoV-2, influenza A virus, influenza B virus and respiratory syncytial virus were detected in our hospital to rule out COVID-19. To detect whether there are interference reaction between the three kind of virus and SARS-CoV-2, RNAs of Influenza A virus (Flu A), Influenza B (Flu B) virus and Respiratory syncytial virus (RSV) were extracted by the same method as SARS-CoV-2, and detected by RT-LAMP and RT-PCR methods respectively.

Furthermore, because we do not have other coronavirus samples and cannot detect whether our primers can amplify other coronaviruses, we compared the primers and target sequences with other virus’ genomic sequences in genebank using BLAST alignment tools.

### Amplification efficiency of RT-LAMP

In order to compare the amplification efficiency of RT-LAMP and RT-PCR, the same RNA was amplified by RT-LAMP and RT-PCR, respectively, and the products were 2-fold serial diluted, and then electrophoresed. Compare the maximum dilution multiples of them.

### Application of RT-LAMP assays on clinical isolates

RT-LAMP detected RNA from a total of 208 nasopharyngeal swab specimens. These specimens have been clinically identified by RT-PCR, of which 17 are SARS-CoV-2 positive, 191 are SARS-CoV-2 negative.

They were from our hospital, Infectious disease hospital of Jinan, and Jinan CDC. Our hospital and Infectious disease hospital of Jinan use the kit of Jiangsu Bioperfectus Technologies Co.,Ltd., while Jinan CDC uses the kit of Shanghai BioGerm Medical Biotedhnology Co.,Ltd..

## Results

### Optimal RT-LAMP Primer for SARS-CoV-2 Detection

We initially screened out one set of optimal RT-LAMP primers for ORF1ab, E and N gene of SARS-CoV-2 respectively. The positions and sequences (Supplemental Table 1) of the optimal primer sets are shown in Figure1.

**Fig. 1.**
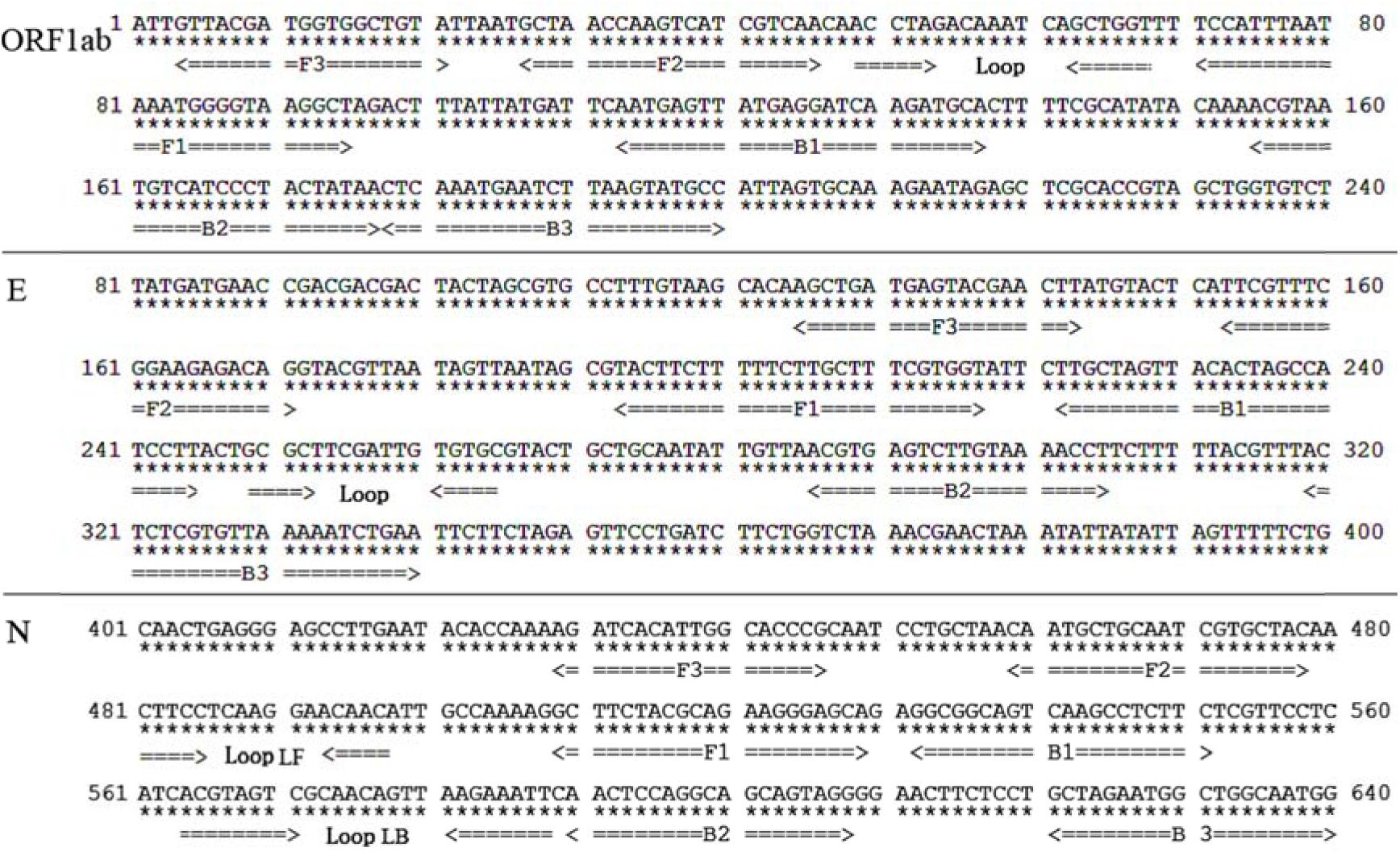
Oligonucleotide primers for SARS-CoV-2 identification. Arrows indicate primer sequences. FIP consists of a complementary sequence of F1 and a sense sequence of F2. BIP consists of a complementary sequence of B1 and a sense sequence of B2. LF/LP primers are composed of the sequences that are complementary to the sequence between F1 and F2 or B1 and B2 regions, respectively.

### Detection of RT-LAMP products

Through Loopamp real-time turbidimeter, typical amplification curves of ORF1ab,E and N gene can be seen. The reaction start time is 32 minutes, 22 minutes, and 24 minutes respectively. The plateau phase is reached within 20 minutes. Figure 2(A) shows the amplification curves of the three genes. Figure 2(B) showed the liquid color of positive reactions changed from orange to green, while negative reactions remained orange. Figure 2(C) showed the product of RT-LAMP was a ladder-like pattern on electrophoresis phase, due to the formation of a mixture of stem-loop DNAs with various stem lengths and cauliflower-like structures with multiple loops formed by annealing between alternately inverted repeats of the target sequence in the same strand.

**Fig. 2.**
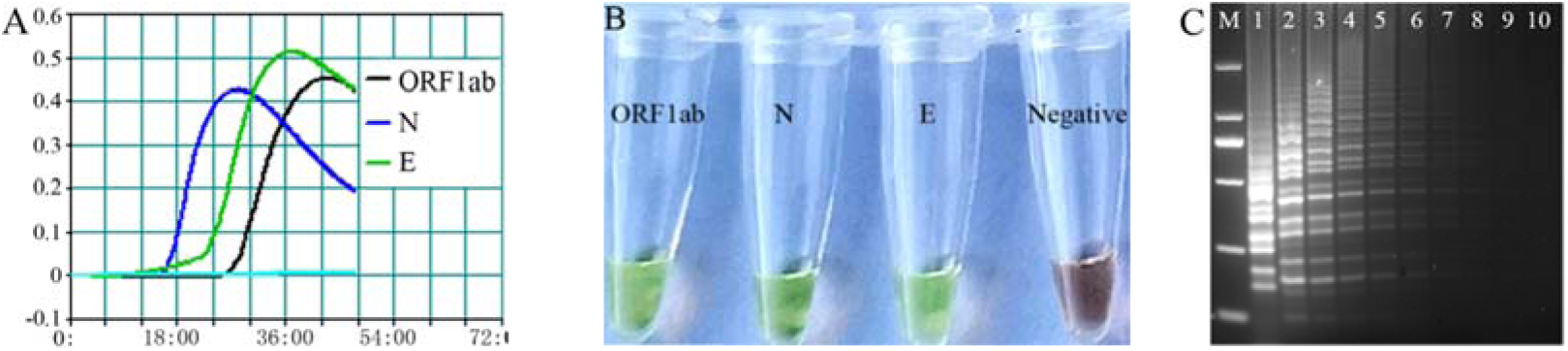
Detection of SARS-CoV-2 by **RT-LAMP** Method. ORF1ab,N and E gene. A: Judge by Loopamp real-time turbidimeter, curves mean positive. B:Judge by naked eyes, green means positive. C: Electrophoresis phase of RT-LAMP products.

### Determination of reaction time

The N gene was positive as early as two samples were positive in 15 minutes, but the false positive of the N gene was also the earliest. The ORF1ab gene was the latest positive. The samples became positive at 20 minutes, and all 5 samples became positive at 30 minutes. However, no false positives occurred until 60 minutes. The E gene is somewhere in between. At 30 minutes, all three genes of the five samples became positive, and 30 minutes can be used as the time point for judgment. The results show that the N gene is the most sensitive, but the specificity is not good. The ORF1ab gene is very specific, but the sensitivity is a little poor. However, by setting a point in time and judging the three combinations, the positive of the sample was determined to be negative (Table 1 and Fig.3.)

**Table 1.**
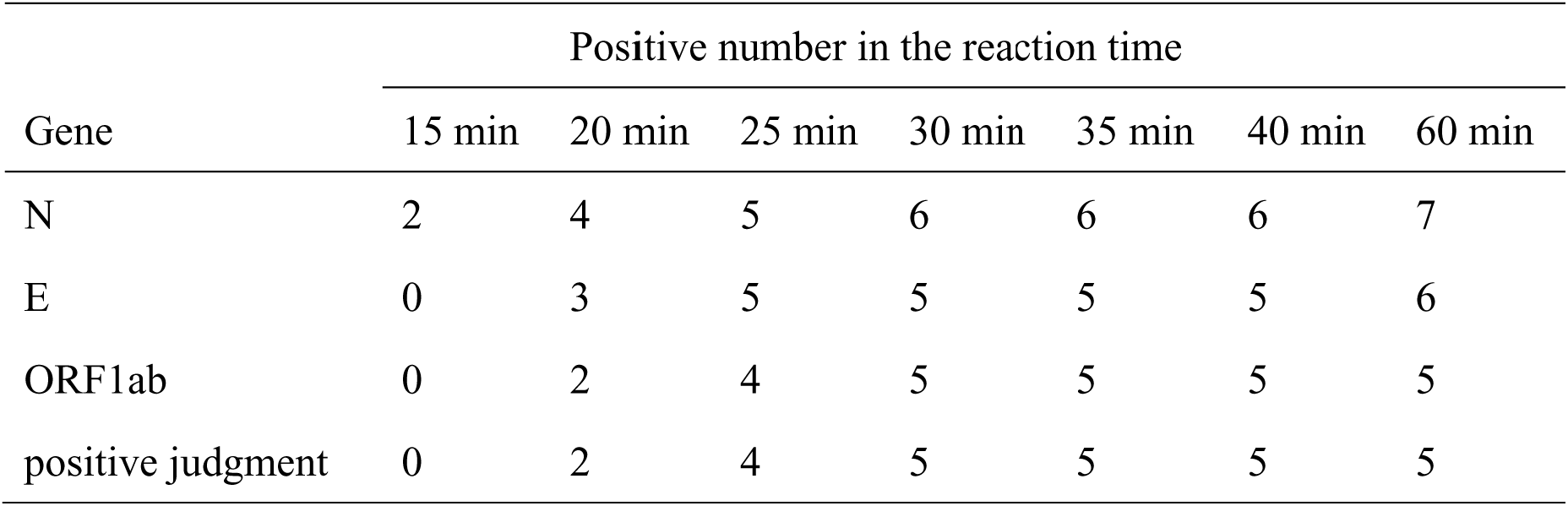
Amplification of ORF1ab, N, E genes for different times by RT-LAMP

| Gene | Positive number in the reaction time |  |  |  |  |  |  |
| --- | --- | --- | --- | --- | --- | --- | --- |
|  | 15 min | 20 min | 25 min | 30 min | 35 min | 40 min | 60 min |
| N | 2 | 4 | 5 | 6 | 6 | 6 | 7 |
| E | 0 | 3 | 5 | 5 | 5 | 5 | 6 |
| ORF1ab | 0 | 2 | 4 | 5 | 5 | 5 | 5 |
| positive judgment | 0 | 2 | 4 | 5 | 5 | 5 | 5 |

**Fig. 3.**
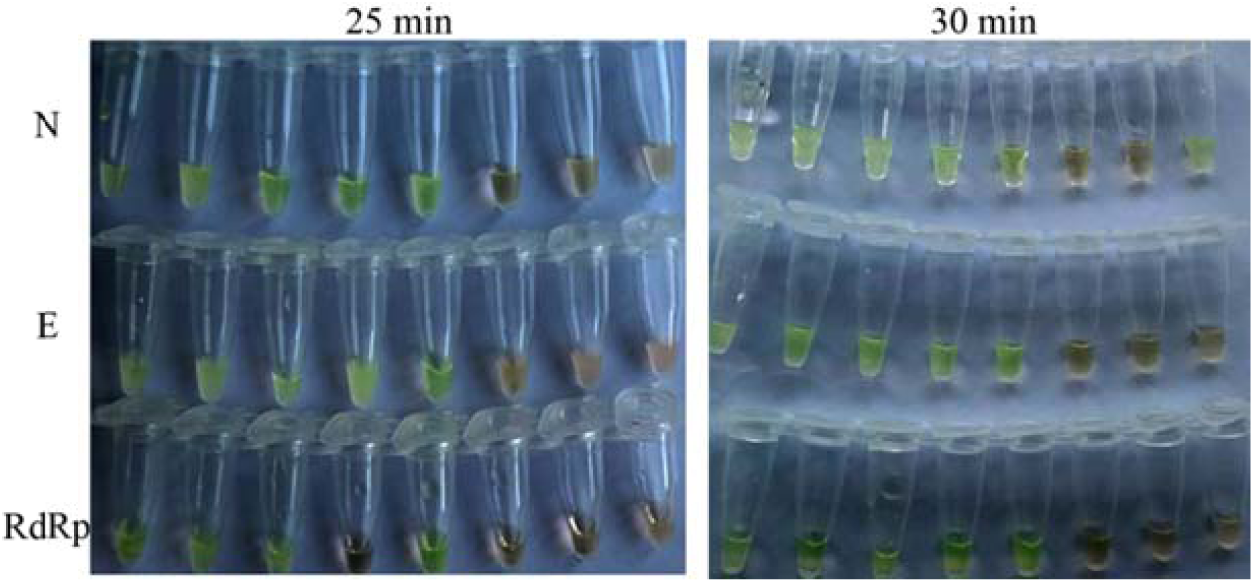
Amplification of ORF1ab, N, E genes at different times by RT-LAMP. After reacting 25 min, N gene and E gene of five samples became positive,and ORF1ab gene of four sample became positive. After reacting 30 min all three genes of the five samples became positive,but there was a false positive for N gene.

### Judgment strategies for SARS-CoV-2 by RT-LAMP

According to the test results, one gene alone is inaccurate to judge the results. In order to obtain more accurate results, the three gene amplification results are combined to judge the existence of SARS-CoV-2. Among them, N is the most sensitive one, detect it can avoid false negatives, while R is the most specific one, detect it can avoid false positives. Table 3 showed the judgment method (Table 2).

**Table 2.** Strategies for judging SARS-CoV-2 positive

| Experiment results | Result |
| --- | --- |
| ORF1ab+ & N + & E+ | SARS-CoV-2 was positive |
| ORF1ab+ & N + / E+ | SARS-CoV-2 was positive |
| ORF1ab + | SARS-CoV-2 was positive |
| N + & E+ | Repeat measurement is required, still N + & E+, SARS-CoV-2 was positive |
| E+ | Repeat measurement is required, when ORF1ab +, SARS-CoV-2 was positive |
| N + | Repeat measurement is required, when ORF1ab + or E+, SARS-CoV-2 was positive |
| ORF1ab- & N- & E- | SARS-CoV-2 was negative |

**Table 3.** Specificity and Interference test (Show positive numbers)

| Samples | N gene | E gene | ORF1ab gene | Positive |
| --- | --- | --- | --- | --- |
| Flu A (10) | 0 | 0 | 0 | 0 |
| Flu B (10) | 1 | 0 | 0 | 0 |
| RSV (10) | 1 | 0 | 0 | 0 |
| Negative (20) | 2 | 0 | 0 | 0 |

### Sensitivity of SARS-CoV-2 identification by the RT-LAMP method

Fig.4 showed that RT-LAMP and RT-PCR have the same sensitivity, and both can detect 20-fold diluted samples. But there are differences for different genes. The N gene can detect a 160-fold diluted sample, the E gene can detect a 40-fold diluted sample, and the ORF1ab gene can detect a 20-fold diluted sample. The detection limit of the RT-PCR kit used is 1000 copIes/ml (5 copies), therefore, the final detection limit of our method is 1000 copIes/ml (5 copies) too.

**Fig. 4.**
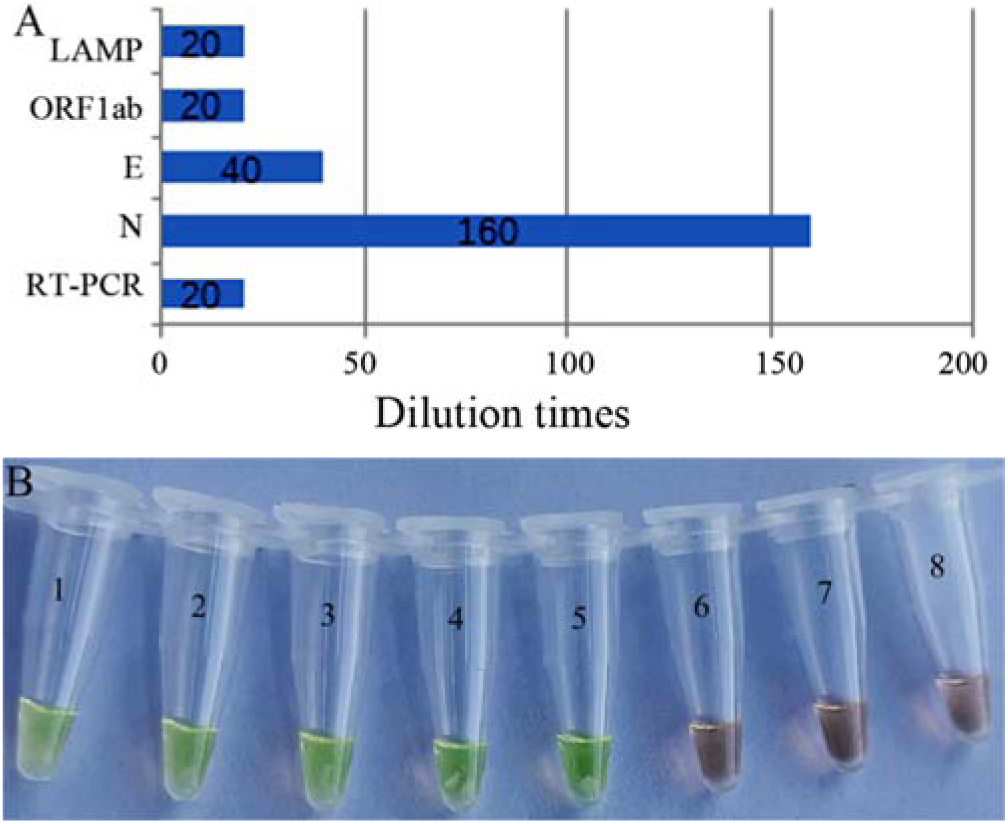
Sensitivity of SARS-CoV-2 identificated by RT-LAMP. A: Comparison of RT-LAMP (three genes) and RT-PCR. N gene can detect a 160-fold diluted sample, the E gene can detect a 40-fold diluted sample, and the ORF1ab gene can detect a 20-fold diluted sample. Combining three genes, RT-LAMP can detect 20-fold diluted samples and with the same sensitivity as PCR. B: Amplification results of N gene after RNA being diluted. 1-8:1,20,40,80,160,320,640,1280 times. N gene can be detected after RNA was diluted for 160 times. Amplification

### Specificity and Interference test of SARS-CoV-2 identification by RT-LAMP

20 SARS-CoV-2 negative samples and 10 Flu A, 10 Flu B and 10 RSV were tested. Table 4 shows that the selected primer sets only amplified the genes of SARS-CoV-2, but could not amplify the genes of Flu A, Flu B and RSV and the negative samples.

The RT-LAMP method using this primer set was specific for SARS-CoV-2 identification.

### Amplification efficiency of RT-LAMP

E gene was amplified by RT-LAMP and RT-PCR, and showed by 3% agarose gel electrophoresis. RT-LAMP product can be observed by electrophoresis after 512 times dilution. However, the RT-PCR product could not be observed by electrophoresis when diluted 512 times, it can be seen when diluted 256 times (Supplemental Figure 1). That is to say, the amplification efficiency of RT-LAMP is higher than that of RT-PCR.

### Application of RT-LAMP assays on clinical isolates

RNA of 208 clinical samples (all were throat and nasal swabs) was detected by RT-LAMP. They had been tested in clinical by RT-PCR, combined with clinical symptoms, 17 of them were confirmed to be positive and 191 were negative. After RT-LAMP amplification for 30 minutes, the results were observed (Fig.5).

**Fig. 5.**
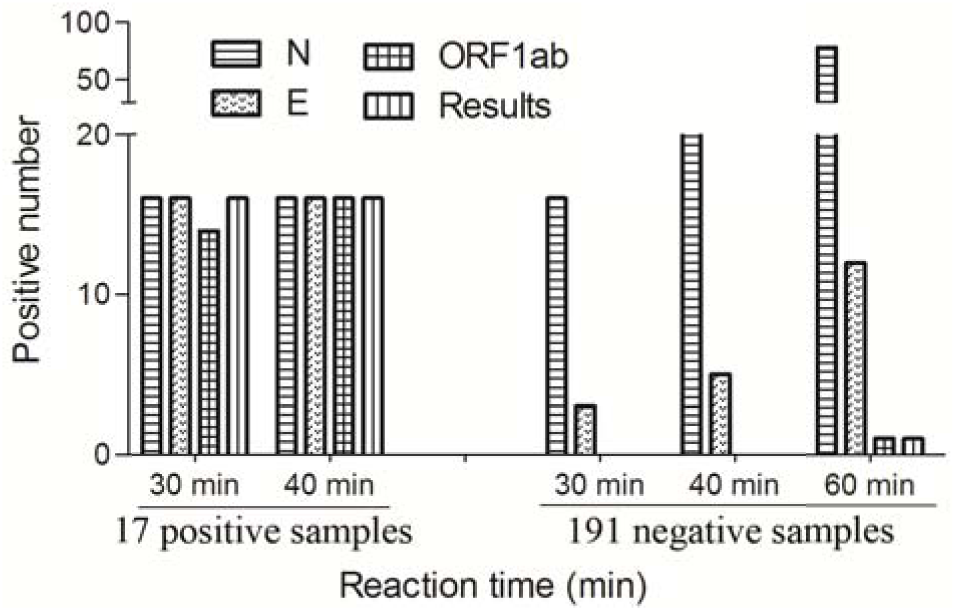
Application of RT-LAMP assays on clinical isolates.

For the 17 positive samples, N gene and E gene of 16 samples were negative, ORF1ab gene of 14 samples was negative. But 10 minutes later, E gene of the other 2 samples was negative. For NO.7 sample, the three genes were all negative. It was tested again by RT-PCR, and it was negative still. The reason should be the RNA had degraded, because it had been stored for a long time and freeze-thaw repeatedly.

For the 191 negative samples, the ORF1ab gene of all the samples was negative, E genes of three samples were negative, and N genes of 16 samples were positive. When judge results after reacting for 40 minutes, E genes of five samples were negative, N genes of 23 samples were positive. An hour after the reaction starting, ORF1ab gene of one sample was positive E gene of 12 samples was positive, and N gene of 78 samples was positive. It is noted that, all three genes of No.1534 sample were positive when judge result at one hour after the reaction starting. These results mean that the results must be observed within the specified time, otherwise false positives will occur. Observe results according to the instructions we have developed, coincidence rate was 99.0 %. As to each gene, coincidence rate of ORF1ab gene, E gene and N gene was 99%, 98.5% and 92.3% respectively. Two samples were positive after 40 minutes of reaction, probably because the RNA concentration was too low, indicating that the extremely low RNA content would lead to false negative.

Of the 17 positive samples, 16 were identified, and one was also negative by RT-PCR. There were two cases in which ORF1ab gene was not detected within the specified time, but because the N gene and E gene were detected, the final judgment was correct. 191 negative samples were judged negative within the specified time. As reacting time increases, false positives appeared. Therefore, the results must be observed at the prescribed time.

## Discussion

COVID-19 is a recently outbreak respiratory disease. It has caused international anxiety due to its rapid spread and high lethality(15). Fast and reliable diagnosis is the top priority for disease control. Now, detecting SARS-CoV-2 nucleic acids by RT-PCR is a clinical standard for the diagnosis of COVID-19.

Although gene amplification methods have been obtained, these methods are time consuming. We developed an RT-LAMP assay to detect triple genes for rapid diagnosis of SARS-CoV-2. The method can complete nucleic acid amplification in 30 minutes at 60-65 °C constant temperature environment. By RT-LAMP, reverse transcription and amplification can be carried out simultaneously useing RNA as template directly. The detection result can be directly judged through the color change by naked eyes, but without needing special instruments. Besides, RT-LAMP analysis is extremely specific because it uses six to eight primers to identify eight different regions on the target DNA. The RT-LAMP assay reported in this paper has many advantages such as operation simpe, reaction fast and detection accurate. The LAMP method has been used to detect hepatitis B virus DNA, Mycobacterium tuberculosis complex, Mycobacterium avian and Mycobacterium intracellular(13, 16).

We designed primers for detecting ORF1ab gene, E gene, and N gene of SARS-CoV-2. About Ten sets of primers were designed for each gene. The optimal primers which can cause specific amplification were selected from these primers. And then loop primers of these optimal primers were designed to shorten the reaction time. Primer design and screening are difficult aspects of LAMP, which takes us a lot of time and energy. After screening out the best primers, we did experiments such as sensitivity, specificity, cross-reaction, and clinical validation.

To compare sensitivity of RT-LAMP and RT-PCR, they were carried out at the same time using the diluted RNA. Results showed that their sensitivity was similar for SARS-CoV-2 judgment. However, for ORF1ab gene, E gene and N gene, RT-LAMP showed different sensitivities. The sensitivity of the N gene is 80 times than the overall sensitivity. Furthermore, products of RT-LAMP and RT-PCR were 2-fold diluted, and were observed by electrophoretic. RT-LAMP is a number of ladder-like bands between 200 bp-2000 bp. Although the RT-PCR kit used also detects three genes, one band appears because the three gene fragments are all about 100 bp. RT-LAMP has one more dilution than RT-PCR, and because there are several bright bands in one electrophoretic lane, it indicates that the amount of RT-LAMP products is very large, which is the reason that the RT-LAMP results are visible by naked eyes. It was verified that influenza A, influenza B, and respiratory syncytial virus-positive specimens did not interfere SARS-CoV-2 detection by RT-LAMP. RNAs extracted from 208 specimens were used for clinical application verification. The 208 specimens had undergone multiple diagnose in clinical such as RT-PCR and clinical symptoms, epidemiological investigations, etc., and had been diagnosed as positive or negative. The accuraty rate of RT-LAMP assay is 99%.

The ORF1ab gene is very specific, the N gene is very sensitive, and the E gene is in between. If using thees genes alone, the accuracy rates of the ORF1ab gene, E gene and N gene are 99%, 98.5%, and 92.3%, respectively, and the accuracy rate of the combined detection results is 99%. SARS-CoV-2 is highly infectious. In order to do not miss suspected cases, we tested the N gene, which is not specific enough but highly sensitive. However, the N gene alone cannot be judged as positive for SARS-CoV-2, and it needs to be re-examined in order to determine with other genes. Through the combined detection of the three genes, the N gene can detect a low concentration of the virus, so that possible positives are not missed, and the ORF1ab gene guarantees that no false positives appear, which also ensures the sensitivity and specificity for SARS-CoV-2 test. However, if one of the three genes is used alone, it can reduce the sensitivity or specificity of the test, causing false positives or false negatives.

In summary, the single-step RT-LAMP measurement reported in this paper is simple, fast, and accurate, and has practicality for SARS-CoV-2 diagnosis in clinical.

## Data Availability

All authors agree that all data submitted here are publicly available.

